# Echocardiography can accurately estimate pulmonary artery wedge pressure without left atrial volume information – diagnostic and prognostic performance

**DOI:** 10.1101/2025.02.04.25321639

**Authors:** Thomas Lindow, Aristomenis Manouras, Geoff Strange, Per Lindqvist, David Playford, Odd Bech-Hanssen, Martin Ugander

**Affiliations:** Department of Pulmonary Medicine, Allergology, and Palliative Medicine, Clinical Sciences, Lund University, Lund, Sweden; Department of Clinical Physiology, Department of Research and Development, Region Kronoberg, Växjö Central Hospital, Växjö, Sweden; Kolling Institute, Royal North Shore Hospital, and University of Sydney, Sydney, Australia; Department of Cardiology, Karolinska University Hospital, and Karolinska Institutet, Stockholm, Sweden; University of Notre Dame, Fremantle, Western Australia, Australia; Department of Clinical Physiology, Umeå University Hospital, Umeå, Sweden; Department of Clinical Physiology, Sahlgrenska University Hospital, Gothenburg, Sweden; Institute of Medicine, Sahlgrenska Academy at the University of Gothenburg, Gothenburg, Sweden; Department of Clinical Physiology, Karolinska University Hospital, and Karolinska Institutet, Stockholm, Sweden

**Keywords:** left ventricular filling pressures, heart failure, echocardiography, left atrium, Doppler

## Abstract

**Background:** A quantitative estimate of pulmonary artery wedge pressures (PAWP) can be obtained using echocardiography, but including left atrial (LA) volume (ePAWP-LA) in the estimation may be misleading. We aimed to derive and validate a new estimate without LA volume information (ePAWP-NOLA) and compare its performance to the ASE/EACVI algorithms for diastolic dysfunction.

**Methods:** ePAWP-NOLA was derived and validated in separate datasets of patients who had undergone right heart catheterization and echocardiography. Prognosis was assessed in the validation cohort and the National Echocardiography Database Australia (NEDA) using Cox regression adjusted for age, sex and left ventricular ejection fraction (LVEF).

**Results:** In the derivation cohort (60±15 years, 40% males, 31% with LVEF<50%), ePAWP-NOLA was derived from mitral (E), and pulmonary vein systolic (S) and diastolic (D) Doppler velocities (n=134, mean difference±SD vs PAWP: 0.0±5.5 mmHg). In the validation cohort (n=116, 51±14 years, 69% males, 89% with LVEF<50%), PAWP agreed with both ePAWP-NOLA and ePAWP-LA (difference 1.3±6.1, 3.2±6.3 mmHg). PAWP>15 mmHg was accurately detected by both ePAWP-NOLA and ePAWP-LA (area under the curve (AUC): 0.84 [0.76–0.92], 0.80 [0.72–0.88]). AUC for the ASE/EACVI algorithm was lower ([0.69 [0.61–0.77]). ePAWP-NOLA and ePAWP-LA correlated with right ventricular afterload, associated with death or implantation of left ventricular assist device (hazard ratio: 1.09 [1.02–1.16], 1.07 [1.02–1.14]), and with cardiovascular death in NEDA (n=38,844, hazard ratio: 1.08 [1.07–1.09], 1.08 [1.07–1.09]).

**Conclusions:** ePAWP-NOLA has diagnostic and prognostic performance comparable to ePAWP-LA, and improved diagnostic performance compared to the ASE/EACVI diastolic dysfunction algorithm.

## Background

Quantification of left ventricular filling pressures is important in detection and grading of heart failure (HF). Although invasive techniques like left- or right-heart catheterization (RHC) offer the most precise determination,(1,2) echocardiography is the preferred bedside method due to its non-invasive nature and widespread accessibility.(3,4) A noninvasive quantitative estimate of pulmonary artery wedge pressure using echocardiography (ePAWP) based on left atrial (LA) volume, mitral peak early velocity (E) and pulmonary vein systolic velocity (S) was recently described.(5) This new measure, ePAWP, showed good agreement with invasively measured PAWP in external validation, high accuracy for detection of elevated PAWP, and was associated with cardiovascular death when applied to a large echocardiographic registry.(5) However, a limitation of the method for estimating ePAWP is its dependency on LA volume. LA remodeling reflects long-term pressure load(6,7) and although related to PAWP, it may not adapt immediately to changes in PAWP. Also, it is associated with interobserver variability due to differences in acquisition or focused or non-focused LA views,(8) and there is a risk of false positive results regarding determining the presence of an elevated PAWP in cases of more prominent LA dilatation or in patients with LA dilatation not caused by increased pressure but arrythmia (atrial fibrillation) or chronic hyperdynamic circulation (anemia, liver failure).(9–11) Therefore, we aimed to derive and validate a quantitative echocardiographic PAWP estimation without LA information (ePAWP-NOLA) in patients with HF symptoms who had undergone echocardiography and right heart catheterization (RHC), to describe their correlation with measures of right ventricular afterload, and compare its performance to ePAWP with LA information (ePAWP-LA) and the current algorithm for diastolic dysfunction. Furthermore, we sought to assess the prognostic strength of these ePAWP measures both in a selected HF population, and in a non-selected, large echocardiographic registry.

## Methods

Four datasets were used in this study, three of them consisting of patients who underwent echocardiography and RHC, the latter based on clinical referral. Two of these datasets, which were used in a previous study to derive and validate ePAWP-LA, together constituted the derivation population for ePAWP-NOLA. The resulting equation was then applied to a third independent dataset for validation regarding agreement with invasive PAWP, change in PAWP (ΔPAWP), and association with death or implantation of left ventricular assist device (LVAD). The prognostic value was further evaluated in the large National Echo Database of Australia (NEDA). Ethical approvals were obtained from the Human Research Ethics Committees for each population, respectively.

### Derivation of ePAWP-NOLA

The training set consisted of patients who had undergone RHC at either Karolinska University Hospital, Stockholm, Sweden (n=153) between 2014 and 2018 or Umeå University Hospital, Umeå, Sweden (n=154) between 2010 and 2015. RHC was performed using a Swan-Ganz thermodilution catheter in the right internal jugular vein, a medial cubital vein, or the right femoral vein. PAWP was defined as the mean PAWP and recorded at end-expirium during spontaneous breathing. Patients with at least moderate mitral valve regurgitation, non-sinus rhythms, constrictive pericarditis, missing PAWP measurements, or missing mitral or pulmonary vein velocities were excluded. The final derivation population consisted of 134 patients for which the main/contributing diagnoses are listed in Supplements, Table S1 and S2. Echocardiographic parameters included in the derivation were mitral E, mitral A, pulmonary vein systolic (S) and diastolic (D) velocities, e’, and relevant variable ratios (E/A, E/e’, S/D, E/S).(5) Tricuspid regurgitation velocity was deliberately not included, in part because of the high prevalence of absent or non-reliable spectral signals during routine echocardiography(12), but in particular due to the association of increased right ventricular systolic pressures and increased pulmonary vascular resistance (PVR) with a broader range of diseases than those causing increased LVFP.(13)

### Validation of ePAWP measures

The validation population consisted of patients who had undergone clinically indicated RHC and echocardiography in close proximity (median [interquartile range] 1 [1–1] days, 90% within 2 days) of the RHC at Sahlgrenska University Hospital, Gothenburg, Sweden (n=275) between 2009 and 2021. The main indications for catheterization were assessment of hemodynamic status as a part of a work-up for heart transplantation, assessment of pulmonary hypertension or restrictive cardiac physiology, and hemodynamic evaluation in cojunction with an endomyocardial biopsy. Patients with non-sinus rhythms (n=61), at least moderate mitral valve regurgitation (n=50), constrictive pericarditis (n=1), missing PAWP measurements (n=1), or missing mitral or pulmonary vein velocities were excluded (n=46).

The final validation population consisted of 116 patients for which the main/contributing diagnoses are listed in Supplements, Table S3. ePAWP-LA was calculated as previously described,(5) ePAWP-LA [mmHg] = 0.179 × LAVI [mL/m^2^] + 2.672 × E [cm/s] / S [cm/s] + 2.7. In addition, ePAWP estimation based on LAVI and mitral E alone, i.e. excluding pulmonary vein velocities, and PAWP estimation based on E/e’ alone were calculated as previously described (ePAWP-E [mmHg] (5) = 0.230 × LAVI [mL/m^2^] + 0.102 × E [cm/s] - 2.7; ePAWP-E/e’ [mmHg](*14*) = 1.24 × E/e’ + 1.9). The results for ePAWP-E and ePAWP-E/e’ are presented as supplemental data.

Besides PAWP, invasive measurements of mean pulmonary artery pressure (mPAP), PVR, pulmonary artery compliance (PAC), calculated as stroke volume divided by pulmonary pulse pressure, pulmonary arterial elastance (Ea), calculated as systolic pulmonary artery pressure divided by stroke volume) were noted for each patient. Through patient records, follow-up was performed until Feb 6, 2024, to obtain survival status or LVAD implantation, which were used as a composite endpoint.

A subset of these patients underwent a repeat RHC and echocardiogram approximately one year later. The difference in ePAWP-NOLA and ePAWP-LA, between the first and second examination is denoted ΔePAWP-NOLA and ΔePAWP-LA.

### Echocardiography

Comprehensive transthoracic echocardiographic exams were performed in all three populations, using a Vivid E9 system (GE Medical Systems, Horten, Norway). Off-line analyses were done using commercially available image analysis software (EchoPAC, General Electric, Waukesha, Wisconsin, USA). Echocardiograms were analyzed by experienced operators blinded to the RHC results. Volumetric and diastolic parameters were obtained and measured according to standard echocardiographic methods.(15) The ASE/EACVI algorithms^6^ for left ventricular filling pressures, based on E/e’, LAVI, and systolic pulmonary artery pressure, were applied to all patients.

### Prognostic performance

Prognostic performance of ePAWP measures was assessed in NEDA, which is a large (>600,000 subjects), observational registry including individual echocardiographic data from participating centers throughout Australia.(16) Typically, included subjects have been referred for echocardiography by a primary care physician in the investigation of known or suspected heart disease, i.e. these patients did *not* exclusively constitute patients with suspected or known HF. By the structure of the Australian health care system, minimal referral bias applies. Data have then been cross-linked to the Australian National Death Index(17) to obtain survival status for each subject until study census date (21 May 2019). In consistency with previous NEDA analyses,(18–20) causes of death were categorized according to ICD-10 and a primary code within I.00–I.99 was considered as a cardiovascular-related death. This classification has been validated previously.(17) The NEDA database has been registered in the Australian New Zeeland Clinical Trials Registry [ACTRN12617001387314].

For each eligible subject, only the last echocardiogram (n=629,721) for each eligible subject in NEDA was included. Patients with biological or mechanical mitral valve prosthesis, pacemaker, mitral stenosis or at least moderate mitral regurgitation, age <18 years, absent follow-up data were excluded (n=98,541). In addition, cases with missing values on LAVI, E, S, D or left ventricular ejection fraction (LVEF) and cases with implausible values (pulmonary venous or mitral velocities <0.005 m/s, LAVI <5 ml/m2) were excluded (n=392,024). A flowchart of the exclusion process is presented in Supplemental Figure S1.

ePAWP-NOLA and ePAWP-LA, were applied, as well as the American Society of Echocardiography (ASE)/European Association of Cardiovascular Imaging (EACVI) algorithms(21) for diastolic dysfunction (algorithm 1) and LA pressure (LAP; algorithm 2), based on the presence of reduced/normal LVEF as recommended.(21) For simplicity, patients with normal diastolic function (ASE/EACVI algorithm 1; LVEF≥50%) and patients with estimated normal LAP (ASE/EACVI algorithm 2; LVEF<50%) were both classified as having normal diastolic function, those with indeterminate results were classified as having indeterminate diastolic function, and those with abnormal results from either algorithm were classified as having abnormal diastolic function.

### Statistical analysis

Data are presented as mean ± standard deviation (SD) or median [interquartile range] as appropriate. Correlations are described using the Pearson correlation coefficient. ePAWP-NOLA was derived by first applying univariable linear regression to echocardiographic parameters that were presumed to be related to PAWP. In a stepwise selection of variables, multivariable linear regression analysis was then performed through different combinations of variables among those with the highest R^2^ in the univariable analysis. Variables were retained in the model only if the adjusted R^2^ increased after addition of a new variable, if a p value <0.05 was obtained when comparing models using the likelihood ratio test, and if the Akaike Information Criteria was reduced by at least 2. The variance inflation factor (VIF) was determined for the variables included in the final model to detect multicollinearity, with low multi-collinearity pre-specified as VIF <3.

The relation between ePAWP-NOLA, and ePAWP-LA with invasively measured PAWP was described using scatterplots in the derivation and validation cohorts, respectively. Diagnostic performance in detection of elevated PAWP (>15 mmHg)(2) was evaluated using receiver operating characteristics (ROC) analysis and described with 95% confidence intervals (95%CI).

In the validation population, the association between ePAWP-NOLA and ePAWP-LA, and the ASE/EACVI diastolic dysfunction grading algorithm respectively, with death or LVAD implantation was evaluated using Cox regression analysis, unadjusted and adjusted for age, sex, and LVEF. A restricted cubic spline regression model was used to demonstrate risk increments with increasing ePAWP-NOLA and ePAWP-LA, respectively. In the NEDA database, the association between ePAWP-NOLA and ePAWP-LA, and the ASE/EACVI diastolic dysfunction grading algorithm respectively, and cardiovascular death was evaluated using Cox regression, unadjusted and adjusted for age, sex, and LVEF. For the purpose of comparison with the categorical values for the ASE/EACVI algorithm, the association with outcomes for ePAWP was also studied after categorizing ePAWP values as follows: ≤15 mmHg, >15–20 mmHg, >20–25 mmHg, and >25 mmHg. As a sensitivity analysis, the association between ePAWP-NOLA, ePAWP-LA and the ASE/EACVI algorithm, respectively, with cardiovascular death was performed after stratification by normal or reduced LVEF and the results are presented in the Supplemental data.

Model discriminatory performance was described with C statistics. Hazard ratios (HR) are presented with 95% confidence intervals (95%CI). To account for the effect of competing risk of death on the analysis using cardiovascular death as an outcome, competing risks regression as described by Fine and Gray was applied to the multivariable analysis and presented with subhazard ratios (SHR) with 95%CI.(22) Statistical significance was accepted at the level of *P* < 0.05 (two-sided). Statistical analysis was performed using R version 4.2.1 (R Core Team, Vienna, Austria).

## Results

### Derivation of ePAWP-NOLA

The derivation population consisted of 134 patients (40 % males, 60±15 years) with a majority of with HF (64.9%) equally distributed between HF with preserved or non-preserved LVEF (34.3 vs 30.6%). Baseline characteristics of the derivation population are described in Table 1. The result of the stepwise linear regression is presented in Table 2. Based on this analysis, the best fit regression equation was ePAWP-NOLA [mmHg] = 2.4 + E [cm/s] / S [cm/s] × 2.76 + D [cm/s] × 0.106. In the derivation cohort, the difference between ePAWP-NOLA and PAWP was 0.0±5.6 mmHg, and correlation was moderate (r=0.64, p<0.001).

**Table 1.**
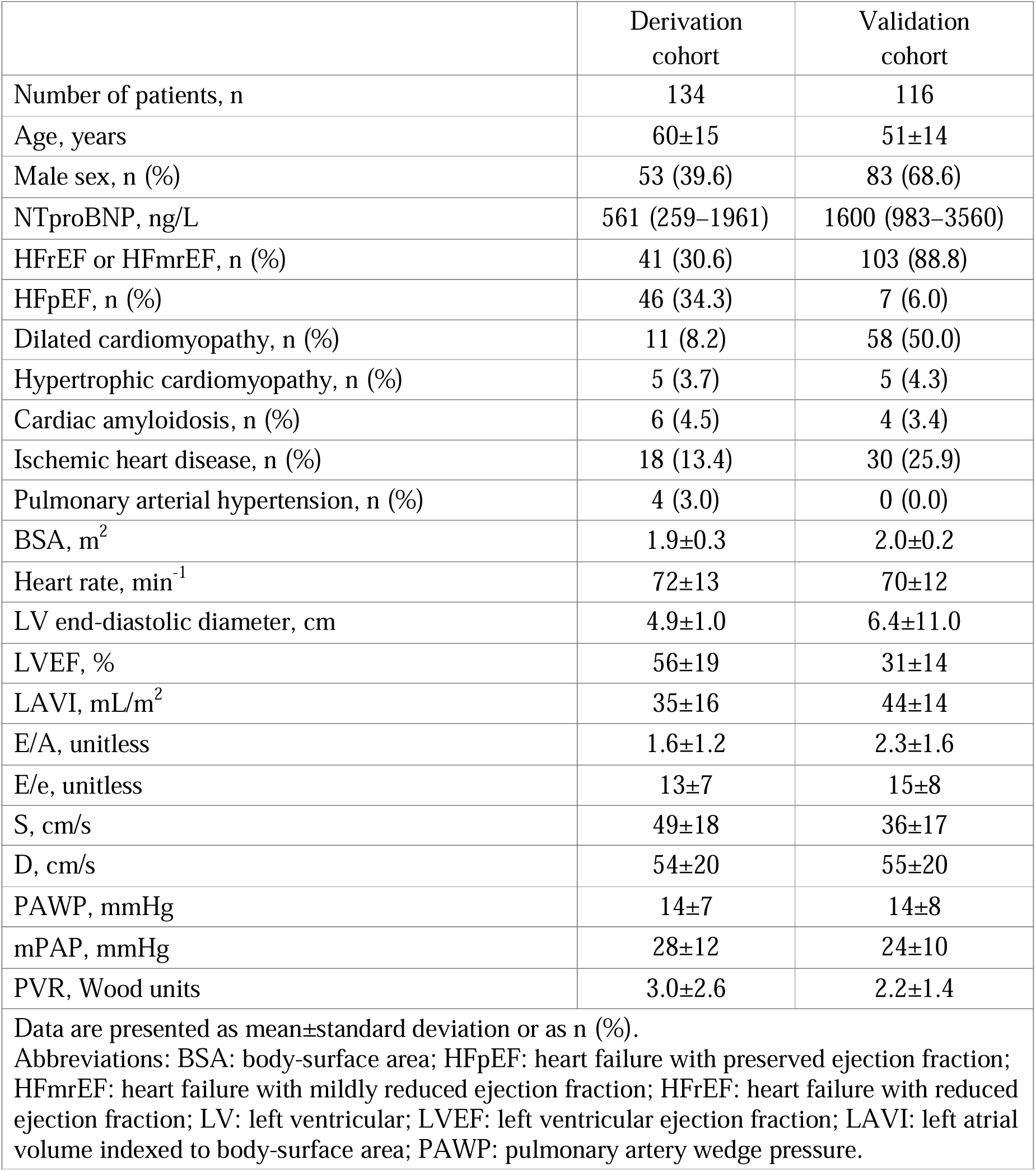
Baseline characteristics for the derivation and validation cohort, respectively.

**Table 2.**
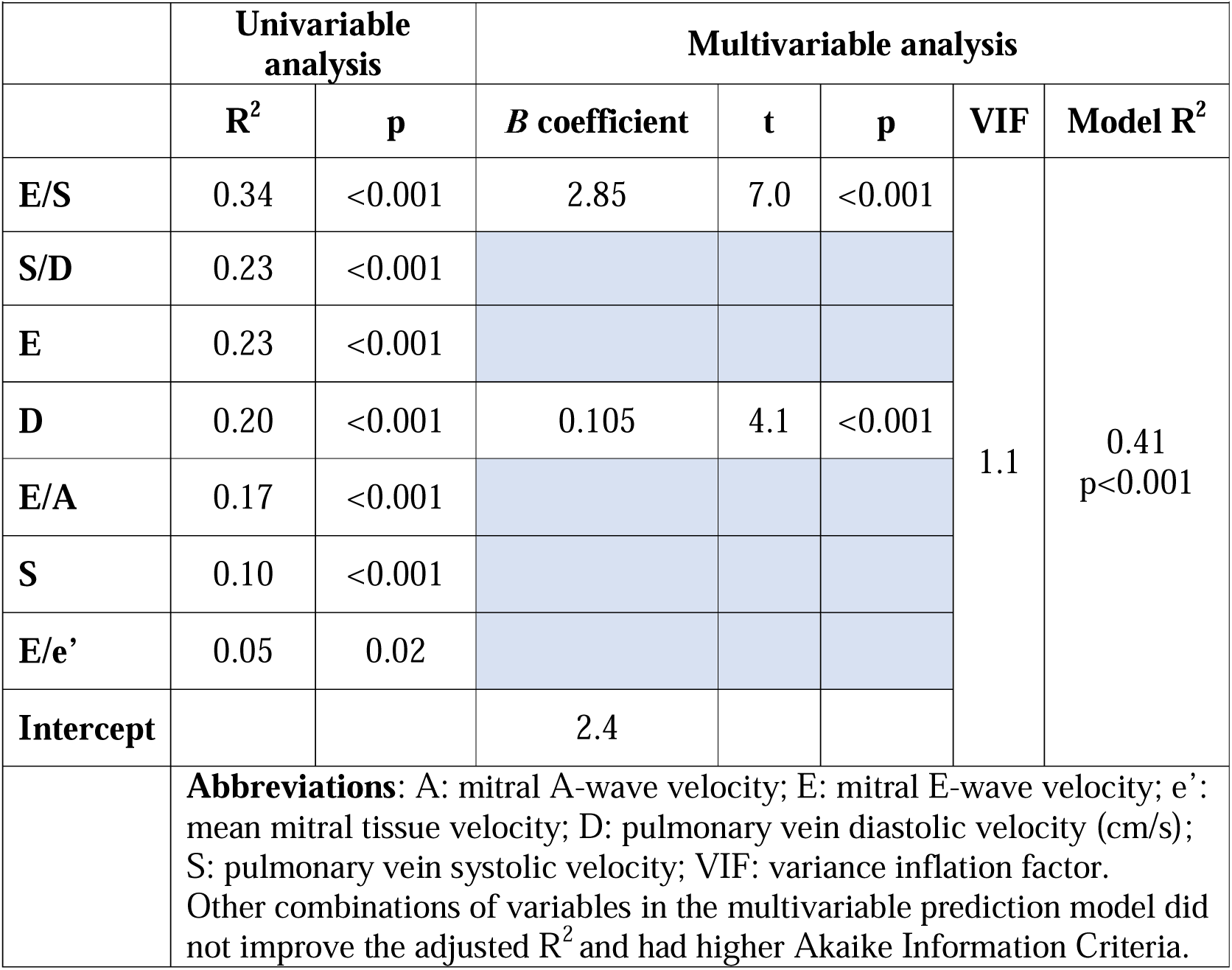
Results of the uni- and multivariable regression analyses for prediction of invasive pulmonary artery wedge pressure using echocardiography in the derivation population (n=134).

### Validation of ePAWP measures

Baseline characteristics of the validation population is presented in Table 1. Compared to the derivation population, patients in the validation population were younger, had more severe left ventricular dysfunction (31±14% compared to 56±19%), higher natriuretic peptide levels and more often had dilated cardiomyopathy or ischemic heart disease. PAWP was similar between the derivation and validation cohorts, while mPAP and PVR were slightly higher in the derivation cohort, see Table 1.

ePAWP-NOLA and ePAWP-LA were applied to 116 patients who had undergone RHC and echocardiography within median (range) 1 (0–6) days (90% within 2 days). Among these patients, ePAWP-NOLA had lower bias (1.3 vs. 3.2 mmHg, p=0.02) but similar precision compared to ePAWP-LA (6.1 vs. 6.3 mmHg, p=0.73), using invasively measured PAWP as reference. Both ePAWP-NOLA and ePAWP-LA correlated with PAWP (r=0.64, r=0.61, p<0.001 for both), and with invasive measures pulmonary artery pressure, right ventricular afterload and NT-proBNP (Table 3). Both ePAWP-NOLA and ePAWP-LA were superior to the ASE/EACVI algorithm for detection of elevated LV filling pressures, Figure 2. ePAWP-NOLA had the highest specificity and positive likelihood ratio, and ePAWP-LA had the highest sensitivity and lowest negative likelihood ratio, Table 4.

**Figure 1.**
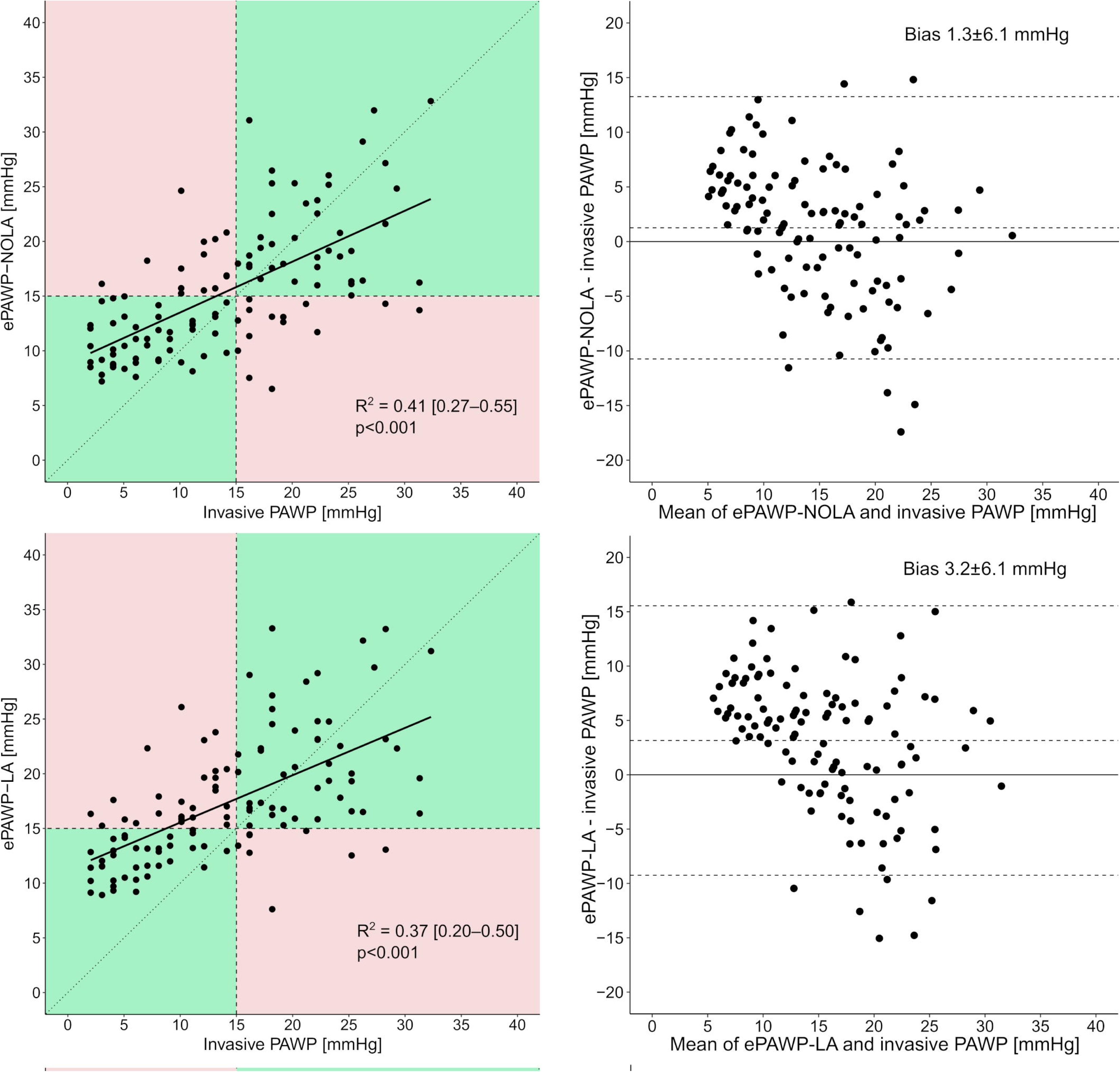
Relation between ePAWP-NOLA (upper panels) and ePAWP-LA (lower panels), with invasively measured pulmonary artery wedge pressure (PAWP) described with scatterplots (left panels) and Bland-Altman plots (right panels) for the validation population (n=116). In the left panels, green boxes denote correct classifications of either normal or elevated invasively measured PAWP defined as ≤ or >15 mmHg, and red boxes denote incorrect classifications.

**Figure 2.**
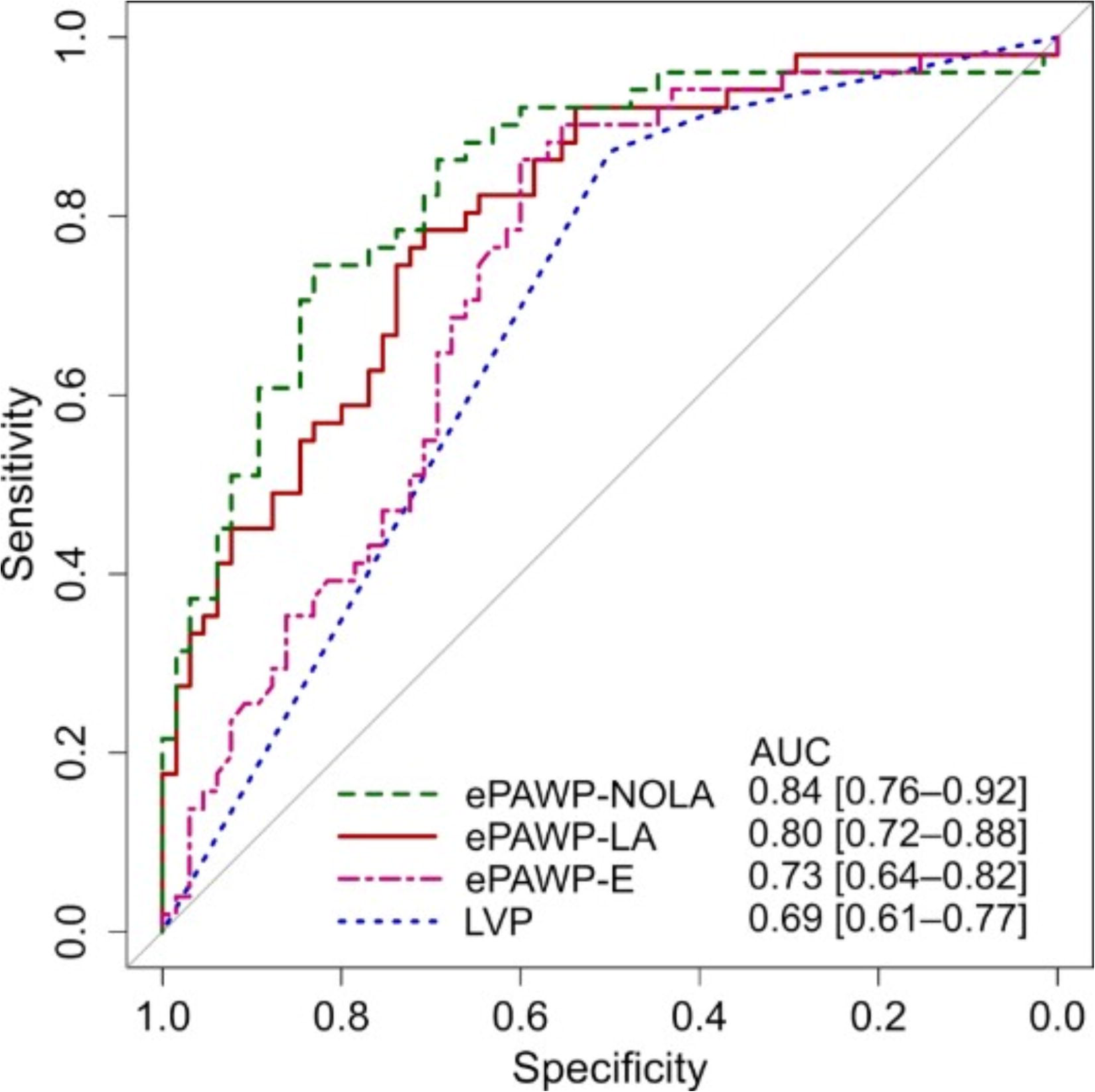
Receiver operating characteristics curve and area under the curve (AUC) with 95% confidence intervals for the detection of elevated pulmonary artery wedge pressure (>15 mmHg) according to right heart catheterization described for ePAWP-NOLA (red solid line), ePAWP-LA (green dashed line), and left ventricular filling pressures according to the ASE/EACVI algorithm (LVP, blue dotted line) in the validation population (n=116).(21)

**Table 3.** Correlation coefficient (Pearson’s r) between pulmonary artery wedge pressure (PAWP), ePAWP-NOLA and ePAWP-LA with invasive hemodynamic measures and NT-proBNP in the validation population (n=116).

|  | PAWP | mPAP | PVR | Ea | PAC | log NT-proBNP |
| --- | --- | --- | --- | --- | --- | --- |
| PAWP | - | 0.88<sup>**</sup> | 0.36<sup>**</sup> | -0.63<sup>**</sup> | 0.78<sup>**</sup> | 0.52<sup>**</sup> |
| ePAWP-NOLA | 0.63<sup>**</sup> | 0.63<sup>**</sup> | 0.38<sup>**</sup> | -0.54<sup>**</sup> | 0.58<sup>**</sup> | 0.40<sup>**</sup> |
| ePAWP-LA | 0.61<sup>**</sup> | 0.61<sup>**</sup> | 0.39<sup>**</sup> | -0.46<sup>**</sup> | 0.56<sup>**</sup> | 0.43<sup>**</sup> |

\* p<0.05, \*\* p<0.001
Abbreviations: mPAP: mean pulmonary arterial pressure; PAC: pulmonary artery compliance; PAWP: pulmonary artery wedge pressure; PVR: pulmonary vascular resistance; Ea: pulmonary arterial elastance.

**Table 4.** Diagnostic accuracy of ePAWP-NOLA, ePAWP-LA and the ASE/EACVI algorithm to detect elevated pulmonary artery wedge pressure (>15 mmHg) according to right heart catherization.

|  | Accuracy | Sensitivity | Specificity | LR+ | ILR- |
| --- | --- | --- | --- | --- | --- |
| ePAWP-NOLA >15 mmHg | 0.79 | 0.75 | 0.83 | 4.4 | 3.3 |
| ePAWP-LA >15 mmHg | 0.71 | 0.86 | 0.58 | 2.1 | 4.2 |
| ASE/EACVI, LVP↑ | 0.67 | 0.87 | 0.50 | 1.7 | 3.8 |
| Abbreviations: LVP: Elevated left ventricular filling pressures according to the ASE/EACVI algorithm(21); LR+: positive likelihood ration; ILR-: inverse negative likelihood ratio (1/negative likelihood ratio) |  |  |  |  |  |

### Detection of change in PAWP

Among 52 patients who performed repeated PAWP and echocardiographic measurements, ePAWP-NOLA could be applied to 24 of them. The repeated examination was performed after 302 [125–818] days. ΔPAWP agreed with ΔePAWP-NOLA and ΔePAWP-LA (−0.6±5.2 and 0.0±5.6 mmHg, respectively) and both ΔePAWP-NOLA and ΔePAWP-LA correlated with ΔPAWP (r=0.82 [0.61–0.91], p<0.001 and r=0.80 [0.59–0.91], p<0.001), Figure 3.

**Figure 3.**
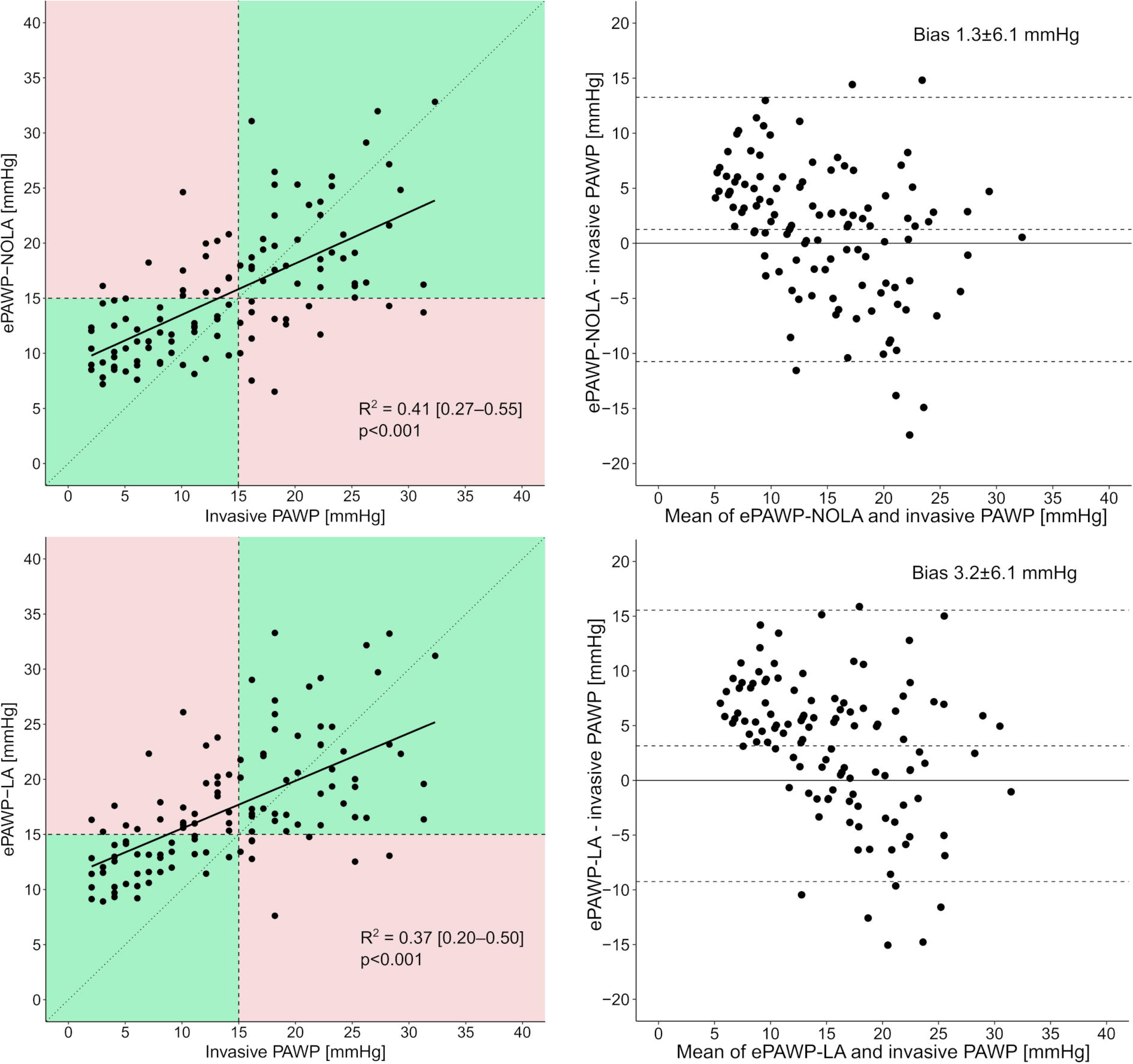
Relation between change (Δ) in ePAWP-NOLA and ΔePAWP-LA and change in invasively measured pulmonary artery wedge pressure (ΔPAWP) described with scatterplots (n=24). In the left panels, green boxes denote correct classifications of the direction of change of invasively measured PAWP, and red boxes denote incorrect classifications.

### Prognostic value of ePAWP measures

During a follow-up of 7 [4–10] years, 36 events occurred (31 deaths). ePAWP-NOLA, and ePAWP-LA were associated with death or LVAD implantation, Table 5. Grading of LV filling pressures according to the ASE/EACVI algorithm was not associated with these outcomes, Table 5. Figure 4 presents the incremental risk increase with increasing ePAWP-NOLA and ePAWP-LA, respectively. For reference, invasively measured PAWP was associated with death or LVAD implantation (HR 1.08 [1.04 - 1.13], adjusted HR 1.08 [1.03 - 1.14]).

**Table 5.** Prognostic value of ePAWP-NOLA, ePAWP-LA and diastolic dysfunction grading according to the ASE/EACVI algorithm presented for both the validation population (n=116, 36 deaths/left ventricular assist implantations) and the NEDA population (n=38,844,2,756 cardiovascular deaths).

|  | Validation cohort |  |  | NEDA cohort |  |  |  |
| --- | --- | --- | --- | --- | --- | --- | --- |
|  | Unadjusted | C | Adjusted | Unadjusted | C | Adjusted | Adjusted SHR |
| ePAWP-NOLA, per 1 mmHg | 1.11 [1.05–1.17] | 0.65 | 1.07 [1.02–1.14] | 1.03 [1.02–1.05] | 0.49 | 1.08 [1.07–1.09] | 1.07 [1.06–1.08] |
| >15-20 mmHg | 1.35 [0.58–3.17] | 0.65 | 1.18 [0.49–2.79] | 1.68 [1.43–1.83] | 0.53 | 2.02 [1.79–2.29] | 1.91 [1.69–2.15] |
| >20-25 mmHg | 3.88 [1.60–9.39] |  | 2.85 [1.10–7.37] | 3.78 [2.79–5.09] |  | 2.12 [1.57–2.87] | 1.91 [1.42–2.59] |
| >25 mmHg | 5.60 [2.09–15.0] |  | 3.41 [1.19–9.83] | 7.55 [4.96–11.5] |  | 3.49 [2.30–5.33] | 2.65 [1.79–3.93] |
| ePAWP-LA, per 1 mmHg | 1.10 [1.04–1.17] | 0.67 | 1.09 [1.02–1.16] | 1.11 [1.10–1.11] | 0.64 | 1.08 [1.07–1.09] | 1.07 [1.07–1.08] |
| >15-20 mmHg | 1.45 [0.61–3.65] | 0.67 | 2.39 [0.91–6.28] | 3.16 [2.89–3.45] | 0.61 | 1.84 [1.68–2.01] | 1.78 [1.62–1.4] |
| >20-25 mmHg | 3.44 [1.35–8.74] |  | 2.40 [0.87–6.62] | 6.53 [5.49–7.77] |  | 2.50 [2.10–2.99] | 2.22 [1.88–2.64] |
| >25 mmHg | 4.22 [1.46–12.18] |  | 4.66 [1.46–14.8] | 9.78 [7.53–12.7] |  | 3.66 [2.80–4.77] | 2.97 [2.24–3.94] |
| ASE/EACVI |  |  |  |  |  |  |  |
| Indeterminate | 0.63 [0.07–5.42] | 0.57 | 0.66 [0.08–5.74] | 3.34 [3.06–3.65] | 0.65 | 1.65 [1.51–1.81] | 1.61 [1.48–1.76] |
| LAP↑ | 2.09 [0.81–5.44] |  | 1.86 [0.71–4.87] | 4.87 [4.41–5.37] |  | 1.83 [1.66–2.03] | 1.80 [1.63–1.99] |

**Abbreviations:** HR: hazard ratio; SHR: subhazard ratio.
The association between ePAWP-NOLA and ePAWP-LA with outcomes are presented both for the continuous ePAWP variable and grouped variables with ePAWP ≤15 mmHg as reference. For the ASE/EACVI algorithm normal diastolic function is the reference.

**Figure 4.**
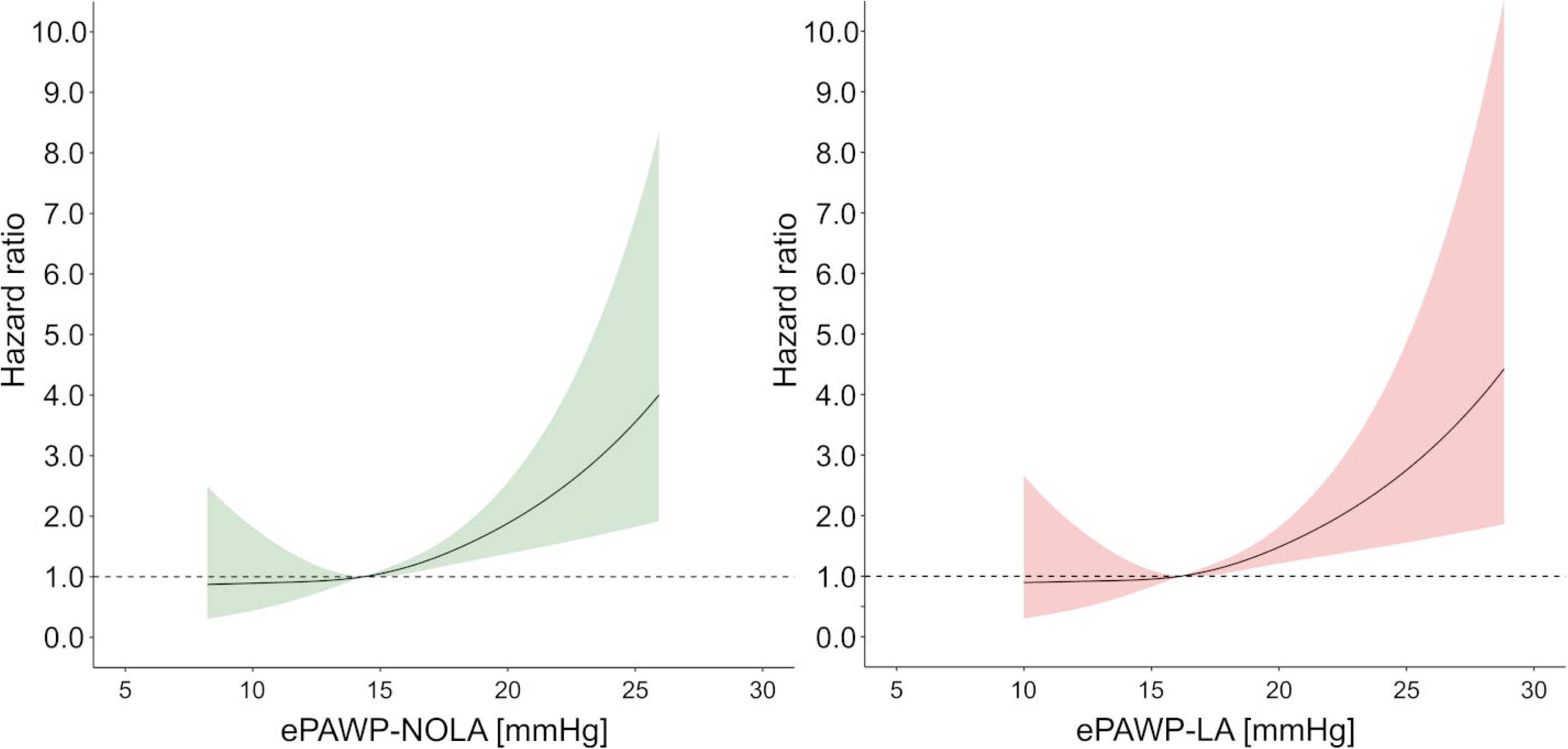
Impact of increasing ePAWP on the risk of death or left ventricular assist implantation for ePAWP-NOLA (left) and ePAWP-LA (right) in the validation population (n=116, follow-up 7 [4–10] years, 36 events). The hazard ratio was calculated using Cox regression and modelled with natural cubic splines with three knots (25th, 50th, and 75th percentiles) for ePAWP-NOLA (left panel) and ePAWP-LA (right panel).

In the NEDA population, ePAWP measures were applied to 38,844 patients with a follow-up of 4.8 [2.3–8.0] years, during which 2,756 cardiovascular deaths occurred. ePAWP measures were associated with cardiovascular death and similar patterns were observed when accounting for competing risks, Table 5. The incremental risk increases with increasing ePAWP values are presented in Figure 5. Diastolic dysfunction grading according to the ASE/EACVI algorithm was also associated with cardiovascular death, in the NEDA population.

**Figure 5.**
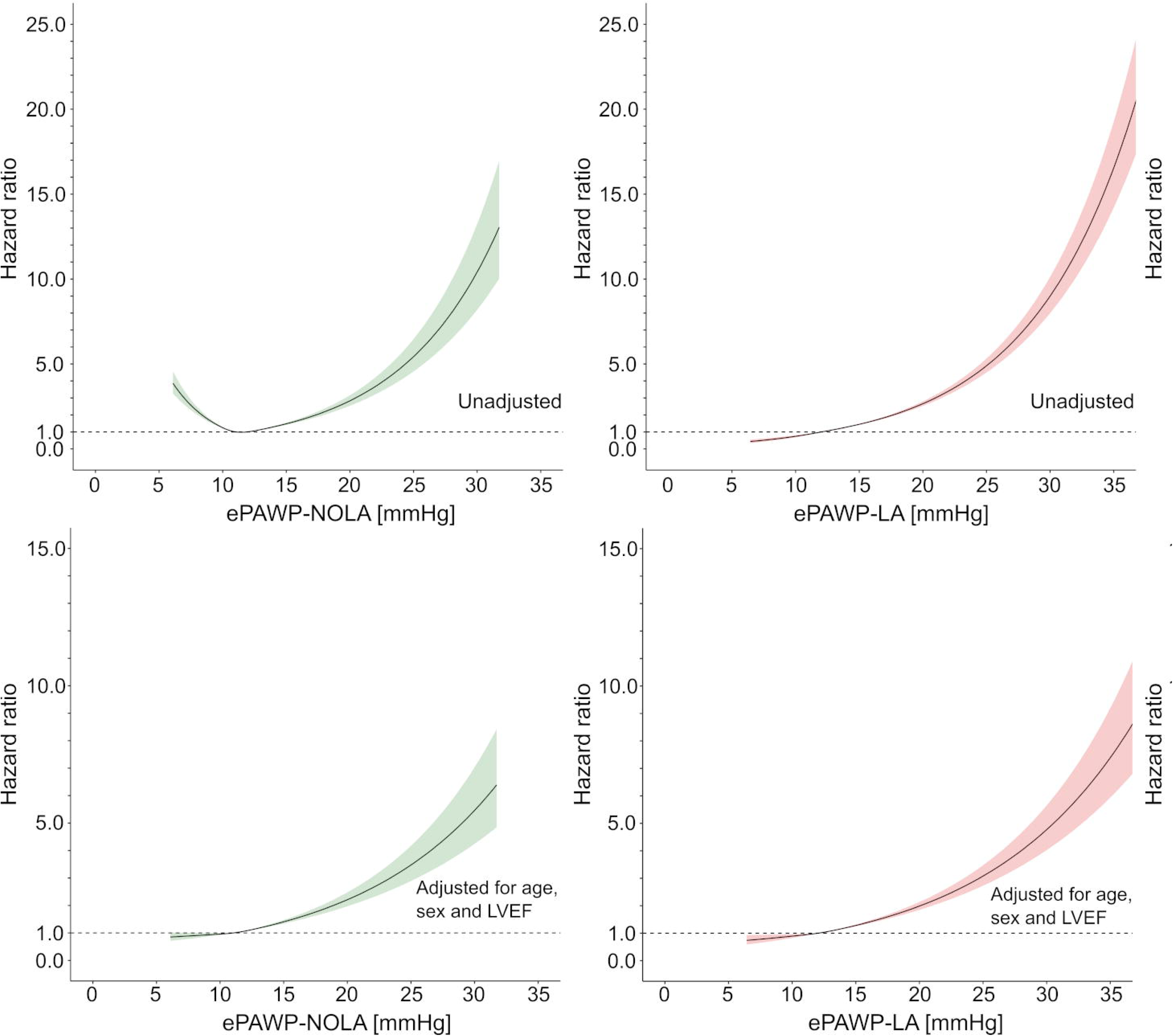
Impact of increasing ePAWP on the risk of cardiovascular death assessed in the National Echocardiography Database of Australia (NEDA), n=38,844, 2,756 cardiovascular deaths, unadjusted (upper panels) and adjusted for age, sex and left ventricular ejection fraction (LVEF) (lower panels). The hazard ratio was calculated using Cox regression and modelled with natural cubic splines with three knots (25th, 50th, and 75th percentiles) for ePAWP-NOLA (left) and ePAWP-LA (right).

Given the increase in HR for ePAWP-NOLA after adjusting for age, the distribution of ePAWP values for ePAWP-NOLA and ePAWP-LA within different age groups is also described in Figure S1. ePAWP-NOLA is higher among the youngest individuals, while ePAWP-LA is evenly distributed between different age groups.

## Discussion

A quantitative echocardiographic estimation of PAWP can be obtained using pulmonary vein velocities (S and D) and early mitral diastolic velocity (E), thereby eliminating the need to include information on LA size. The regression equation can easily be incorporated into echocardiographic software for automatic and immediate presentation. ePAWP-NOLA derived in patients with echocardiography and RHC showed acceptable accuracy in prediction of PAWP, good ability to detect elevated PAWP and was associated with incrementally increasing risk of death or LVAD implantation among patients with HF. Change in ePAWP using both ePAWP-NOLA and ePAWP-LA showed strong correlation with change in invasively measured PAWP. This emphasizes the clinical use of a quantitative PAWP estimate, since algorithmic solutions only can provide information on whether left ventricular filling pressures are elevated or not.

When compared to invasively measured PAWP, ePAWP-NOLA showed a numerically lower bias (1.5 mmHg) compared to ePAWP-LA (3.2 mmHg) in PAWP estimation, which contrasts previous reports of low bias for ePAWP-LA vs. PAWP.(5) However, the current validation cohort consisted of a larger proportion of patients with left heart disease, and HF with reduced ejection fraction (HFrEF) in particular. Relying on LA volume in PAWP estimation would require a linear relation between change in PAWP and change in LA volume, which may not be the case since chronic LA dilatation may remain even in the presence of a reduction in LV filling pressure.(23) Conversely, from a prognostic perspective, chronic LA dilatation acting as a surrogate for persistently elevated LV filling pressure, is strongly associated with a higher mortality. Thus, consideration of both current (ePAWP) and long-term (LAVI) LV filling pressure offers helpful clinical information.

Both ePAWP-NOLA and ePAWP-LA had higher diagnostic accuracy than the ASE/EACVI algorithm for detection of elevated PAWP. For ePAWP-NOLA improvement in specificity was substantial. The superior diagnostic accuracy is in line with previous ePAWP reports(5), although the AUC was lower in this study (ePAWP-NOLA 0.84, ePAWP 0.80) compared to the original ePAWP validation populations (0.94).(5) Compared to the original validation population, the current study consisted of a larger proportion of patients with HFrEF and larger LA volumes. In addition, a quantitative estimate on a continuous scale provides information on severity of PAWP elevation with greater granularity than the three classes obtained with the ASE/EACVI algorithm.Pulmonary vein velocities are important determinants of both ePAWP-NOLA and ePAWP-LA. The relation between PAWP and pulmonary vein velocities has been described previously.(24–26) The components of the S wave are affected by both LA relaxation, which in turn is affected by mitral annular plane systolic excursion (MAPSE), (27,28) right ventricular function, and LA compliance.(25) Although the S wave is composed of two different waveforms, these are fused to one in the vast majority of cases evaluated by transthoracic echocardiography.(29) The D-wave velocity, occurring during LA conduit phase, is affected by the same factors that determine the transmitral pressure gradient as reflected by the mitral E, such as the elastic recoil of the LV and LV stiffness, which will affect PAWP.(30) The mitral E, and the pulmonary D, is also affected by loading conditions, mitral valve impedance, and blood viscosity.(31) Given the close relationship between E and D, it could be presumed that D would carry the same diagnostic information as E, and E/S the same information as S/D. However, E/S was more strongly associated with PAWP than S/D. Moreover, D but not E provided incremental value to E/S when determining the regression equation.

Doppler interrogation of pulmonary vein velocities has been described to be difficult, and is not recommended in routine assessment of LV filling pressures.(21) Other studies suggest pulmonary vein velocity acquisitions to be highly feasible,(32) in particular when included in standard clinical protocols(5), suggesting that routine exclusion of these measurements is not particularly justified.

Both ePAWP-NOLA and ePAWP-LA correlated with invasive measures of increased RV pulsatile afterload (PAC). This strengthens the pathophysiological rationale of including these measures in the assessment of LV filling pressures. An increase in RV afterload is expected in these patients both by backward transmission of increased LA pressure but also as a long-term remodelling effect on pulmonary vasculature. Both increased resistance and reduced capacitance of the pulmonary vasculature are likely to contribute to reduced S-wave velocities, in particular.(24,29)

We showed a moderate correlation between ePAWP-NOLA and PAWP, and ePAWP-LA and PAWP, respectively. This is in line with the original description of ePAWP,(5) but contrasts another recent report.(33) Prediction of increased PAWP by echocardiography based on the ASE/EACVI algorithm compared to cardiovascular magnetic resonance imaging (CMR) using LA volumes and LV mass has shown a higher accuracy for echocardiography than for CMR.(33) In a subset of patients in that study, ePAWP could be determined and showed weak correlation (r=0.34). Bias was not reported, and the number of patients included was small (n=32). Also, no information on the variation of PAWP in that subset was provided.

All ePAWP measures are limited by their precision in reference to invasive PAWP measurements. Of note, even invasive measurements are afflicted by variation at repeated examinations (±2.1 mmHg).(34) This affects its robustness as a reference standard, e.g. when deriving a quantitative echocardiographic measure, and also puts the observed precision of ePAWP measures in perspective. In that sense, simultaneous measurements of invasive and echocardiographic PAWP would be preferred, but was performed only in a subset of the derivation population (the Umeå cohort). In the Karolinska population, and in the validation cohort echocardiography was performed in close proximity in time to the right heart catheterization.

The strong accuracy provided by ePAWP-NOLA in reference to PAWP, makes this estimation suitable for PAWP estimation in larger studies, allowing for comparisons of PAWP between groups. Also, given the strong correlation between change in ePAWP measurements and change in PAWP suggests utility for using ePAWP to assess treatment effects in larger studies.

All ePAWP measurements were associated with increased risk of death or LVAD implantation, while the ASE/EACVI algorithm was not. In the validation cohort, similarly strong associations were found for invasively measured PAWP. Increasing ePAWP-NOLA and ePAWP-LA were all associated with increasingly higher risk, in line with a previous study of ePAWP.(5) The HR for ePAWP-NOLA was higher after adjusting for age, sex and left ventricular ejection fraction. While mitral E and D are higher among younger individuals, S is usually lower. Thus, age is likely to confound the association between ePAWP-NOLA and cardiovascular death with higher ePAWP-NOLA values being more common among the youngest, often healthier individuals. This was confirmed by excluding patients younger than 30 years from the analysis, after which there was an unadjusted association between ePAWP-NOLA and cardiovascular death. In the HF population (the validation population), a strong association with outcomes was found for ePAWP-NOLA even in the unadjusted analysis. This is important, since it suggests that ePAWP-NOLA is robust and provides prognostically relevant information when applied to patients with suspected HF, and ePAWP results must be interpreted in the context of clinical information.

### Limitations

Our findings are limited in part by small sample sizes, and clinically heterogenous composition of the derivation and validation populations. The adequate performance of ePAWP-NOLA in populations with variable proportions of HFrEF can also be considered a strength allowing for wider application of these estimations.

The evaluation of the prognostic value of ePAWP-NOLA, ePAWP-LA and diastolic dysfunction in the NEDA database was performed after numerically substantial exclusions, which is a limitation. However, the association with outcomes was found also in the validation cohort, which strengthen the finding of an association between these measures and adverse outcomes.

Therefore, these measures need to be validated in prospective, clinical settings in patients with suspected or confirmed HF.

Also, possible differences in echocardiographic acquisition and the absence of a core study laboratory are limitations of our study. Both LA volume and Doppler measurements are dependent on acquisition.(8) However, our results reflect real world settings. Neither patients with moderate mitral valve lesions, pacemaker, nor with atrial fibrillation were included in this study, and ePAWP estimations cannot, therefore, be applied to such patients. Future attempts to derive quantitative PAWP estimations in these patients is merited.

## Conclusion

Compared to ePAWP-LA, ePAWP-NOLA provides comparable diagnostic and prognostic performance, and improved diagnostic performance compared to current diastolic dysfunction guideline algorithms.

## Funding

This work has been funded in part by grants to TL from Region Kronoberg, to MU from New South Wales Health, Heart Research Australia, and the University of Sydney, to PL from The Swedish Heart and Lung foundation (20160787, 20200160) and the Swedish Research Council (2019–01338).

## Author contributions

Study design: TL, MU. Data acquisition: AM, PL, GS, DP, PL, OBH. Data analysis: TL, AM, GS, DP, PL, OBH, MU. Data interpretation: All. Drafting the manuscript: TL, MU. Critical revision of the manuscript for important intellectual content: All. Agreement to be accountable for the work: All.

## Conflicts of interest

**TL:** none**; AM:** none**; GS:** Consulting fees from NEDA**; PL:** None**; DP**: Consulting fees from NEDA**; OBH:** None**; MU:** none.

## Data availability

The data underlying this article can be shared upon reasonable request to the corresponding author, except for the data obtained through the National Echocardiography Database of Australia.

## Abbreviations

E: Mitral early peak wave velocity
HF: Heart failure
LAVI: Left atrial volume indexed to body surface area
LAP: Left atrial pressure
LVEF: Left ventricular ejection fraction
LVFP: Left ventricular filling pressures
PAWP: Pulmonary artery wedge pressure
S: Systolic pulmonary vein velocity
D: Diastolic pulmonary vein velocity
RHC: Right heart catheterization

## Notes

### Funding Statement

This work has been funded in part by grants to TL from Region Kronoberg, Sweden, to MU from New South Wales Health, Heart Research Australia, and the University of Sydney, to PL from The Swedish Heart and Lung foundation (20160787, 20200160) and the Swedish Research Council (2019-01338).

### Author Declarations

For the invasive cohorts, ethical approval was obtained from the Swedish Ethical Review Authority and informed consent was obtained from study participants. For the National Echocardiographic Database of Australia, ethical approval was obtained from the Royal Prince Alfred Human Research Ethics Committe, who waived informed consent.

## References

1. Kubiak GM, Ciarka A, Biniecka M, Ceranowicz P. Right Heart Catheterization-Background, Physiological Basics, and Clinical Implications. J Clin Med 2019;8:1331.

2. Humbert M, Kovacs G, Hoeper MM et al. 2022 ESC/ERS Guidelines for the diagnosis and treatment of pulmonary hypertension. Eur Resp J 2023;61.

3. Gardin JM, Leifer ES, Fleg JL et al. Relationship of Doppler-Echocardiographic left ventricular diastolic function to exercise performance in systolic heart failure: the HF-ACTION study. Am Heart J 2009;158:S45–52.

4. Terzi S, Sayar N, Bilsel T et al. Tissue Doppler imaging adds incremental value in predicting exercise capacity in patients with congestive heart failure. Heart Vessels 2007;22:237–44.

5. Lindow T, Manouras A, Lindqvist P et al. Echocardiographic estimation of pulmonary artery wedge pressure - invasive derivation, validation, and prognostic association beyond diastolic dysfunction grading. Eur Heart J Cardiovasc J 2024;25:498–509.

6. Douglas PS. The left atrium: a biomarker of chronic diastolic dysfunction and cardiovascular disease risk. J Am Coll Cardiol 2003;42:1206–7.

7. Hammoudi N, Achkar M, Laveau F et al. Left atrial volume predicts abnormal exercise left ventricular filling pressure. Eur J Heart Fail 2014;16:1089–95.

8. Kebed K, Kruse E, Addetia K et al. Atrial-focused views improve the accuracy of two-dimensional echocardiographic measurements of the left and right atrial volumes: a contribution to the increase in normal values in the guidelines update. Int J Cardiovasc Imaging 2017;33:209–218.

9. Lang RM, Badano LP, Mor-Avi V et al. Recommendations for cardiac chamber quantification by echocardiography in adults: an update from the American Society of Echocardiography and the European Association of Cardiovascular Imaging. J Am Soc Echocardiogr 2015;28:1–39.e14.

10. Kaur H, Premkumar M. Diagnosis and Management of Cirrhotic Cardiomyopathy. J Clin Exp Hepatol 2022;12:186–199.

11. Shafi I, Harmouch KM, Prakash P, Kahe F, Ramappa P, Afonso L. Unmasking High-Output Heart Failure: Beyond Conventional Paradigms. Cardiol Rev 2025.

12. McQuillan BM, Picard MH, Leavitt M, Weyman AE. Clinical correlates and reference intervals for pulmonary artery systolic pressure among echocardiographically normal subjects. Circulation 2001;104:2797–802.

13. Rosenkranz S, Gibbs JS, Wachter R, De Marco T, Vonk-Noordegraaf A, Vachiéry JL. Left ventricular heart failure and pulmonary hypertension. Eur Heart J 2016;37:942–54.

14. Nagueh SF, Middleton KJ, Kopelen HA, Zoghbi WA, Quiñones MA. Doppler tissue imaging: a noninvasive technique for evaluation of left ventricular relaxation and estimation of filling pressures. J Am Coll Cardiol 1997;30:1527–33.

15. Mitchell C, Rahko PS, Blauwet LA et al. Guidelines for Performing a Comprehensive Transthoracic Echocardiographic Examination in Adults: Recommendations from the American Society of Echocardiography. J Am Soc Echocardiogr 2019;32:1–64.

16. Strange G, Celermajer DS, Marwick T et al. The National Echocardiography Database Australia (NEDA): Rationale and methodology. American heart journal 2018;204:186–189.

17. Magliano D, Liew D, Pater H et al. Accuracy of the Australian National Death Index: comparison with adjudicated fatal outcomes among Australian participants in the Long-term Intervention with Pravastatin in Ischaemic Disease (LIPID) study. Aust N Z J Public Health 2003;27:649–53.

18. Playford D, Strange G, Celermajer DS et al. Diastolic dysfunction and mortality in 436L360 men and women: the National Echo Database Australia (NEDA). Eur Heart J Cardiovasc Imaging 2021;22:505–515.

19. Strange G, Stewart S, Celermajer D et al. Poor Long-Term Survival in Patients With Moderate Aortic Stenosis. J Am Coll Cardiol 2019;74:1851–1863.

20. Strange G, Stewart S, Celermajer DS et al. Threshold of Pulmonary Hypertension Associated With Increased Mortality. J Am Coll Cardiol 2019;73:2660–2672.

21. Nagueh SF, Smiseth OA, Appleton CP et al. Recommendations for the Evaluation of Left Ventricular Diastolic Function by Echocardiography: An Update from the American Society of Echocardiography and the European Association of Cardiovascular Imaging. Eur Heart J Cardiovasc Imagin 2016;17:1321–1360.

22. Fine JP, Gray RJ. A Proportional Hazards Model for the Subdistribution of a Competing Risk. J Am Stat Assoc 1999;94:496–509.

23. Thomas L, Marwick TH, Popescu BA, Donal E, Badano LP. Left Atrial Structure and Function, and Left Ventricular Diastolic Dysfunction: JACC State-of-the-Art Review. J Am Coll Cardiol 2019;73:1961–1977.

24. Appleton CP. Hemodynamic determinants of Doppler pulmonary venous flow velocity components: new insights from studies in lightly sedated normal dogs. J Am Coll Cardiol 1997;30:1562–74.

25. Smiseth OA, Thompson CR, Lohavanichbutr K et al. The pulmonary venous systolic flow pulse--its origin and relationship to left atrial pressure. J Am Coll Cardiol 1999;34:802–9.

26. Rossvoll O, Hatle LK. Pulmonary venous flow velocities recorded by transthoracic Doppler ultrasound: relation to left ventricular diastolic pressures. J Am Coll Cardiol 1993;21:1687–96.

27. Keren G, Sherez J, Megidish R, Levitt B, Laniado S. Pulmonary venous flow pattern--its relationship to cardiac dynamics. A pulsed Doppler echocardiographic study. Circulation 1985;71:1105–12.

28. Keren G, Sonnenblick EH, LeJemtel TH. Mitral anulus motion. Relation to pulmonary venous and transmitral flows in normal subjects and in patients with dilated cardiomyopathy. Circulation 1988;78:621–9.

29. Fadel BM, Pibarot P, Kazzi BE et al. Spectral Doppler Interrogation of the Pulmonary Veins for the Diagnosis of Cardiac Disorders: A Comprehensive Review. J Am Soc Echocardiogr 2021;34:223–236.

30. Tabata T, Thomas James D, Klein Allan L. Pulmonary venous flow by doppler echocardiography: revisited 12 years later. J Am Coll Cardiol 2003;41:1243–1250.

31. Smiseth OA, Thompson CR. Atrioventricular filling dynamics, diastolic function and dysfunction. Heart Fail Rev 2000;5:291–9.

32. Jensen JL, Williams FE, Beilby BJ et al. Feasibility of obtaining pulmonary venous flow velocity in cardiac patients using transthoracic pulsed wave Doppler technique. J Am Soc Echocardiogr 1997;10:60–6.

33. Rahi W, Hussain I, Quinones MA, Zoghbi WA, Shah DJ, Nagueh SF. Noninvasive Prediction of Pulmonary Capillary Wedge Pressure in Patients With Normal Left Ventricular Ejection Fraction: Comparison of Cardiac Magnetic Resonance With Comprehensive Echocardiography. J Am Soc Echocardiogr 2024;37:486–494.

34. Melillo CA, Lane JE, Aulak KS, Almoushref A, Dweik RA, Tonelli AR. Repeatability of Pulmonary Pressure Measurements in Patients with Pulmonary Hypertension. Ann Am Thorac Soc 2020;17:1028–1030.

