## Supplemental data for "Echocardiography can accurately estimate pulmonary artery wedge pressure without left atrial volume information – diagnostic and prognostic performance"

**SUPPLEMENTS**

| **Table S1. Principal and/or contributing diagnoses after right heart catheterization in the Karolinska cohort (part of the derivation cohort)** | |
| --- | --- |
| Heart failure with preserved ejection fraction | 17 (18.9) |
| Heart failure with reduced/mildly reduced ejection fraction | 10 (11.1) |
| Ischemic heart disease | 9 (10) |
| Pre-/post heart transplantation | 6 (6.7) |
| Restrictive cardiomyopathy | 5 (5.6) |
| Right ventricular failure | 5 (5.6) |
| Pulmonary fibrosis | 5 (5.6) |
| Normal | 4 (4.4) |
| Aortic stenosis | 4 (4.4) |
| Chronic thromboembolic pulmonary hypertension | 4 (4.4) |
| Systemic sclerosis | 4 (4.4) |
| Dilated cardiomyopathy | 3 (3.3) |
| Cardiac amyloid | 3 (3.3) |
| Hypertrophic cardiomyopathy | 3 (3.3) |
| Myocarditis | 2 (2.2) |
| Pulmonary hypertension | 2 (2.2) |
| Chronic obstructive pulmonary disease | 2 (2.2) |
| Arythmogenic cardiomyopathy | 1 (1.1) |
| Cirrhosis | 1 (1.1) |
| Idiopathic pulmonary arterial hypertension | 1 (1.1) |
| Respiratory insufficiency | 1 (1.1) |

| **Table S2. Principal and/or contributing diagnoses after right heart catheterization in the Umeå cohort (part of the derivation cohort)** | |
| --- | --- |
| Heart failure with reduced/mildly reduced ejection fraction | 11 (20.8) |
| Systemic sclerosis | 6 (11.3) |
| Normal | 5 (9.4) |
| Heart failure with preserved ejection fraction | 5 (9.4) |
| Associated pulmonary arterial hypertension | 5 (9.4) |
| Chronic thromboembolic pulmonary hypertension | 5 (9.4) |
| Systemic lupus erythematosus /Mixed connective tissue disease | 4 (7.5) |
| Idiopathic pulmonary arterial hypertension | 3 (5.7) |
| Ischemic heart disease | 2 (3.8) |
| Chronic obstructive pulmonary disease | 2 (3.8) |
| Hypertrophic cardiomyopathy | 1 (1.9) |
| Aortic stenosis | 1 (1.9) |
| Ventricular septum defect | 1 (1.9) |
| Cardiac amyloid | 1 (1.9) |

| **Table S3. Principal and/or contributing diagnoses after right heart catheterization in the validation cohort** | |
| --- | --- |
| Myocarditis | 4 (3.4) |
| Amyloidosis | 4 (3.4) |
| Aortic regurgitation | 3 (2.6) |
| Dilated cardiomyopathy | 57 (49.1) |
| Peripartal cardiomyopathy | 1 (0.9) |
| Hypertrophic cardiomyopathy | 5 (4.3) |
| HFpEF | 1 (0.9) |
| Hypertension | 1 (0.9) |
| Ischemic heart disease | 30 (25.9) |
| Unspecified | 4 (3.4) |
| Myositis | 2 (1.7) |
| Sarcoidosis | 2 (1.7) |
| Restrictive cardiomyopathy | 2 (1.7) |

**Regression equation for estimation of pulmonary arterial wedge pressure (PAWP) using only left atrial volume indexed to body surface area (LAVi) and mitral early peak velocity (E) or E/e’**

*ePAWP_E =_ 0.230 × LAVi + 10.177 × mitral E - 2.7,* in which ePAWP is given in mmHg, LAVi in ml/m^2^ and mitral E in m/s

*ePAWP-E/e’ [mmHg]^11^ = 1.24 × E/e’ + 1.9*

*Comparison of PAWP estimation ePAWP-E and ePAWP-E/e’*

ePAWP-NOLA, ePAWP-LA, ePAWP-E and ePAWP-E/e’ could be applied to the same 79 patients in the validation cohort. In this subset, ePAWP-E/e’ had larger bias and worse precision (6.5±11 mmHg) compared to ePAPW-NOLA (1.3±6.1 mmHg), ePAWP-LA (3.2±6.4 mmHg) and ePAWP-E (0.1±6.7). In contrast to the other ePAWP measures, ePAWP-E/e’ was not associated with outcomes in the validation population, Table S3.

In the NEDA population, the necessary measures to apply both ePAWP-NOLA, ePAWP-LA, ePAWP-E and ePAWP-E/e’ were available in 18,342 patients, which incurred 1,548 cardiovascular deaths. In this subset, all four ePAWP measures were associated with cardiovascular death after adjusting for age, sex and LVEF. ePAWP-NOLA was not associated with cardiovascular death in the univariable analysis, Table 4. In a post-hoc explorative analysis, there was an association between ePAWP-NOLA and cardiovascular death after excluding patients younger than 30 years (HR 1.05 [1.03–1.07]).

| **Table S4. Comparative overview of quantitative echocardiographic methods for the estimation of pulmonary artery wedge pressure (PAWP) regarding agreement with invasive reference and prognosis (cardiovascular mortality) in patients in which all required measurements for ePAWP and E/e’ were available (LAVI, E, S, D, e’).** | | | | | | | |
| --- | --- | --- | --- | --- | --- | --- | --- |
| **Method** | **Formula** | **Difference vs invasive PAWP [mmHg]** | **AUC [95%CI]**  **for PAWP**  **>15 mmHg** | **Hazard ratio**  (95% CI)^‡^ | | | |
|  |  |  |  | Validation population  (n=116, 36 events) | | NEDA population  (n=18,342, 1,548 events) | |
|  |  |  |  | Unadjusted | Adjusted for age, sex, and LVEF | Unadjusted | Adjusted for age, sex, and LVEF |
| ePAWP-NOLA | E/S × 2.76 + D × 0.106 + 2.4 | 1.5±6.2 | 0.83 [0.74–0.93] | 1.09 [1.02 – 1.15] | 1.07 [1.01–1.14] | 1.00 [0.98–1.02]^†^ | 1.08 [1.06–1.10] |
| ePAWP-LA | LAVI × 0.179  + E/S × 2.672 + 2.7 | 3.3±6.3 | 0.80 [0.70–0.90] | 1.10 [1.03–1.17] | 1.11 [1.03–1.19] | 1.11 [1.10–1.11] | 1.08 [1.07–1.10] |
| ePAWP-E | LAVI × 0.230  + E × 0.102 - 2.7 | 0.1±6.7 | 0.73 [0.61–0.84] | 1.11 [0.99–1.25] | 1.08 [0.98–1.19] | 1.08 [1.07–1.09] | 1.06 [1.05–1.08] |
| E/e’^11^ | E/e' × 1.24 + 1.9 | 6.5±11 | 0.63 [0.51–0.75] | 0.98 [0.96–1.04] | 1.00 [0.96–1.04] | 1.09 [1.08–1.09] | 1.04 [1.04–1.05] |
| **Abbreviations**: AUC: area under the curve; CI: confidence interval; LAVI: left atrial volume indexed to body surface area (ml/m^2^); E: mitral early peak wave velocity (cm/s); S: systolic peak pulmonary vein velocity (cm/s); LVEF: left ventricular ejection fraction (%); D: diastolic peak pulmonary vein velocity (cm/s).  † In a post-hoc explorative analysis, there was an association between ePAWP-NOLA and cardiovascular death after excluding patients younger than 30 years (HR 1.05 [1.03–1.07]). See text for details. | | | | | | | |

¨

| **Table S5.** Prognostic value of ePAWP-NOLA, ePAWP-LA and diastolic dysfunction grading according to the ASE/EACVI algorithm stratified by left ventricular ejection fraction (LVEF) in the NEDA population | | | | | | |
| --- | --- | --- | --- | --- | --- | --- |
|  | LVEF<50%  (n=35,488, 2,131 cardiovascular deaths) | | | LVEF>50%  (n=3,356, 625 cardiovascular deaths) | | |
|  | HR, unadjusted | C | HR, adjusted | HR, unadjusted | C | HR, adjusted |
| ePAWP-NOLA,  per 1 mmHg | 1.00 [0.99–1.02] | 0.47 | 1.08 [1.06–1.09] | 1.08 [1.05–1.10] | 0.58 | 1.09 [1.07–1.11] |
| ePAWP-LA, per 1 mmHg | 1.10 [1.10–1.11] | 0.62 | 1.08 [1.07–1.09] | 1.12 [1.10–1.13] | 0.66 | 1.09 [1.08–1.11] |
| ASE/EACVI |  |  |  |  |  |  |
| Indeterminate | 3.33 [3.01–3.69] | 0.64 | 1.57 [1.42–1.75] | 1.61 [1.34–1.94] | 0.57 | 1.68 [1.40–2.02] |
| Diastolic dysfunction/LAP↑ | 5.31 [4.74–5.95] |  | 1.89 [1.68–2.12] | 1.56 [1.28–1.91] |  | 1.50 [1.22–1.83] |
| **Abbreviations**: HR: hazard ratio; LAP: left atrial pressure | | | | | | |

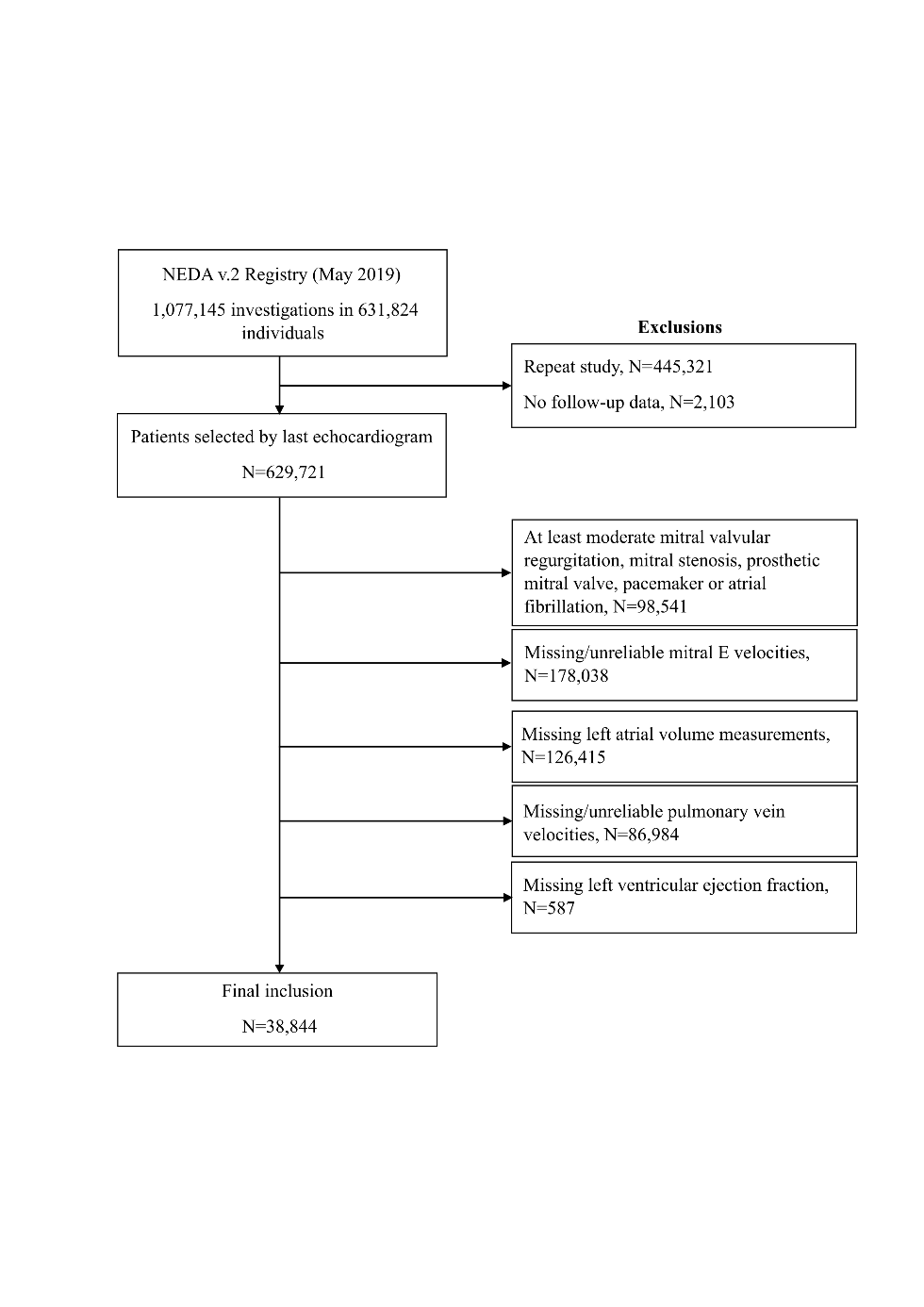

**Figure S1.** Flowchart of patient inclusion and exclusion in the NEDA database.

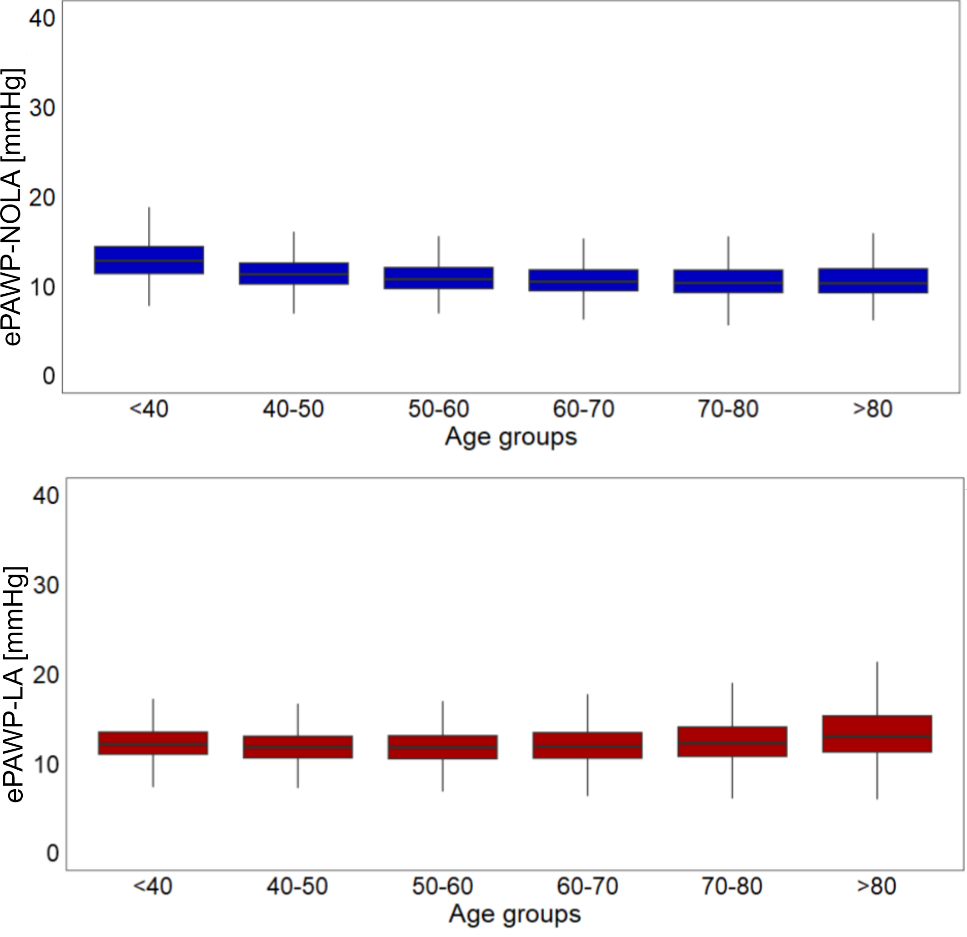

**Figure S2.** Distribution ePAWP-NOLA (upper panel) and ePAWP-LA (lower panel) stratified by age among patients in the NEDA population. Notably, ePAWP-NOLA are highest for younger individuals.
